# Investigating the genetic pathways of insomnia in Autism Spectrum Disorder

**DOI:** 10.1101/2022.02.03.22270340

**Authors:** Maria Niarchou, Emily V. Singer, Peter Straub, Beth A. Malow, Lea K. Davis

## Abstract

**Background:** Sleep problems are common in children with autism spectrum disorder (autism). There is sparse research to date to examine whether insomnia in people with autism is related to autism genetics or insomnia genetics. Moreover, there is a lack of research examining whether circadian-rhythm related genes share potential pathways with autism.

**Aims:** To address this research gap, we tested whether polygenic scores of insomnia or autism are related to risk of insomnia in people with autism, and to test whether the circadian genes are associated with insomnia in people with autism.

**Methods and procedures:** We tested these questions using the phenotypically and genotypically rich MSSNG dataset (N=1,049) as well as incorporating in the analyses data from the Vanderbilt University Biobank (BioVU) (N=349).

**Outcomes and results:** In our meta-analyzed sample, there was no evidence of associations between the polygenic scores (PGS) for insomnia and a clinical diagnosis of insomnia, or between the PGS of autism and insomnia. We also did not find evidence of a greater burden of rare and disruptive variation in the melatonin and circadian genes in individuals with autism and insomnia compared to individuals with autism without insomnia.

**Conclusions and implications:** Overall, we did not find evidence for strong effects of genetic scores influencing sleep in people with autism, however, we cannot rule out the possibility that smaller genetic effects may play a role in sleep problems. Our study indicated the need for a larger collection of data on sleep problems and sleep quality among people with autism.

## Introduction

Sleep problems have been reported in 44 to 86% of children with Autism Spectrum Disorder (autism) as compared to 10 to 16% of typically developing (TD) children, with insomnia (difficulty falling asleep, staying asleep, short sleep duration, or early morning awakening) presenting as the most common sleep problem^1^. A study on 1193 children with autism found that children with sleep problems had more internalizing and externalizing behavior problems and worse adaptive skill development than children with autism without sleep problems^2^. In another study of 2714 children with autism, core symptoms of autism, including social impairment and restricted/repetitive behaviors, were associated with short sleep duration^3^. Clinical and animal studies have shown that sleep problems during development are associated with long-lasting developmental consequences that can affect learning, memory and mood regulation^4^. Importantly, sleep disturbance can have profound negative impacts on children with autism and their families^5, 6^.

Despite the importance and frequency of sleep disturbance in the autism population, the mechanisms underlying risk of sleep problems in autism remain poorly understood and relatively understudied. Recent genome-wide association studies (GWAS), have shown that hundreds of genetic variants with small effects are likely to contribute to risk for autism ^7^ and insomnia^8^. Furthermore, insomnia in the general population is heritable and influenced by many genetic risk factors^9^. Although individual loci have small effects on risk, multi-locus approaches have been used to enhance the information provided by GWAS. The polygenic risk profiling approach provides a composite measure of genetic risk and involves a weighted sum of even moderately associated alleles on a GWAS chip into a single genetic score for each individual. This genetic score is subsequently used to predict case status (e.g., presence or absence of a diagnosis) in independent datasets. Though imperfect, this method has shown that autism genetic profile risk scores (PRS) can partly distinguish autism cases from controls. Insomnia PRS similarly distinguish those with insomnia from controls in the general population^8^. However, there is no research to date examining the overlap in autism and insomnia genetic architecture or examining whether insomnia PRS could be used to predict insomnia in individuals with autism.

Melatonin is an important physiological sleep regulator, as it helps regulate the circadian rhythms and synchronize our sleep-wake cycle^10^. Controversy also exists regarding melatonin levels in individuals with autism. Some studies have found lower levels of overnight or morning blood melatonin levels and in the enzyme involved in melatonin production (ASMT) in individuals with autism compared to those with TD (e.g.,^11^) while a small study documented normal overnight blood melatonin profiles, including in children whose insomnia responded to supplemental melatonin (e.g.,^12, 13^). Another study documented a relationship between melatonin-related pathways genes and sleep-onset delay^14^. Additionally, two small studies reported associations between clock genes and autism ^15, 16^. A study by Nicholas and colleagues (2007)^15^, screened 11 clock/clock-related genes in 110 individuals with autism and their parents. A significant association was found for Per1 and Npas2. Although, given the small sample size, the associations did not withstand correction for multiple testing^15^, the association was replicated by Yang and colleagues (2016)^16^ who analysed 28 autism patients (14 with sleep disorders and 14 without) and 23 control subjects of Japanese descent. Finally, another study identified two nonsense, one frameshift and one de novo missense PER2 variants in four individuals with autism out of 5,100 (0.08%)^17^. Of these four, three reported a history of sleep problems and one had symptoms of familial advanced sleep-phase syndrome 1. However, a study on 2,065 children with autism did not identify any associations between variation in expression of circadian genes and sleep problems^18^. The associations between clock genes and autism indicate potential shared pathways, but further research in larger samples is essential to replicate and clarify these associations.

The aims of our study were to examine (1) the genetic contribution of insomnia polygenic scores (PGS) established in the typically developed population, to risk of insomnia in individuals with autism, and (2) the extent to which melatonin and circadian-relevant genes and pathways are associated with insomnia in individuals with autism.

## Materials and methods

The project was approved by the Vanderbilt University Medical Center (VUMC) Institutional Review Board (IRB #190564).

We used data from two sample collections, namely BioVU and MSSNG (see descriptions below).

### BioVU

BioVU is Vanderbilt’s biobank that stores DNA samples from over 250,000 individuals that comes from discarded blood and is linked to the clinical data from the Synthetic Derivative (SD)^19^. The SD is a mirror de-identified version of the VUMC Electronic Health Records (EHR), including over 3 million records with a wealth of information (e.g., diagnostic and procedure codes (ICD 9/ICD 10 and CPT), basic demographics (age, sex, race), text from clinical care including discharge summaries, nursing notes, progress notes, history and physicals, problem lists and multi-disciplinary assessments, laboratory values, clinical text and electronically derived trace values, and inpatient medication orders).

### MSSNG

The MSSNG dataset (https://research.mss.ng) is a unique resource that comes from an initiative to collect genetic samples that have been whole genome sequenced (WGS) as well as phenotypic information from individuals with autism and their families, and make this available to the research community. It is a joint effort of Autism Speaks, University of Toronto, SickKids Hospital, and Google and is the largest collection of available WGS for researchers. For a description of the MSSNG infrastructure see here^20, 21^.

### Genotyping and quality control

#### BioVU sample

The genotyping of the participants that were in our sample was completed on the Illumina MEGA^EX^ platform that includes over two million genetic markers. Details on the procedures of quality controls are described elsewhere^22^. We performed genotype imputation using the Michigan Imputation Server ^23^, while as a reference panel we used the Haplotype Reference Consortium (HRC) reference panel. All variants with a low imputation quality (i.e., R^2^ <0.3) were removed. Cryptic relatedness was addressed by removing one individual of each pair for which pihat>0.2. To define individuals of primarily European ancestry we used FlashPCA2 combined with CEU, YRI, and CHB reference sets from the 1000 genomes project phase 3 spiked in and the Europeans were selected. The cut-off for the European subset was within 30% of the CEU cluster on the CEU-YRI axis and within 40% of the CEU cluster on the CEU-CHB axis.

#### MSSNG sample

Variant call files were downloaded from the MSSNG (version db6). We filtered the genotype data for variants with a MAF<0.01, removed SNPs with more than 10% missing data, minimum read depth of 0.10 and maximum reading depth of 75). We further selected individuals of European ancestry based on the MSSNG database attribution that was a consensus of reported ancestry (where available), admixture analysis and an inference model based on this method^24^. To select unrelated individuals, after filtering for individuals with phenotypic data (see process below), we also removed individuals with a pi-hat of >0.2. To derive the rs numbers, we merged the MSSNG filtered data with the 1000 European Genomes build 38. A total of 6,241,225 variants on autosomes across 1,049 individuals were available to be tested. The top 10 principal components were calculated using flashpca2. For the SNP-set (Sequence) Kernel Association Test (SKAT) analysis (see Statistical analyses below), we followed the same filtering process, without removing variants based on their MAF.

### Phenotypes

#### BioVU

Individuals with autism were identified in the SD using an algorithm that was created and validated by clinician experts (for a description of the process see:^25^). ES and MN, under the supervision of BM (an expert in sleep issues in autism), completed a comprehensive chart review of all individuals with genotype data and autism in BioVU (n=349) to assess whether the patient had sleep problems or not. Sleep problems were defined as insomnia-related problems and were defined as either sleep onset insomnia (more than 30 minutes to fall asleep), sleep maintenance insomnia (waking up through the night and difficulty falling back to sleep) or short sleep duration (less than 6 hours of sleep per night). Further details of our process are described here^25^.

#### MSSNG

Cases were diagnosed with autism if they met criteria based on scores from standardized diagnostic criteria tools, typically the Autism Diagnostic Observation Schedule (ADOS), and the Autism Diagnostic Interview-Revised (ADI-R) and/or was supported by clinical criteria. The definition of sleep problems in the MSSNG data is described in Figure 1. Specifically, following the definition of insomnia in BioVU, we defined sleep problems in MSSNG as any problem with sleep initiation, sleep maintenance and short sleep duration. In the case of medical history records, if the participant was assessed as having a sleep disorder or any sleep problem, this would also qualify as a sleep problem. In terms of frequency, we only counted a sleep problem as such if they occurred sometimes, or more often. We did not include as sleep problems a range of other sleep-related issues (e.g., restless sleep, body pains during sleep, sleep anxiety, sleep disordered breathing) as their relationship to insomnia genes are unclear.

**Figure 1.**
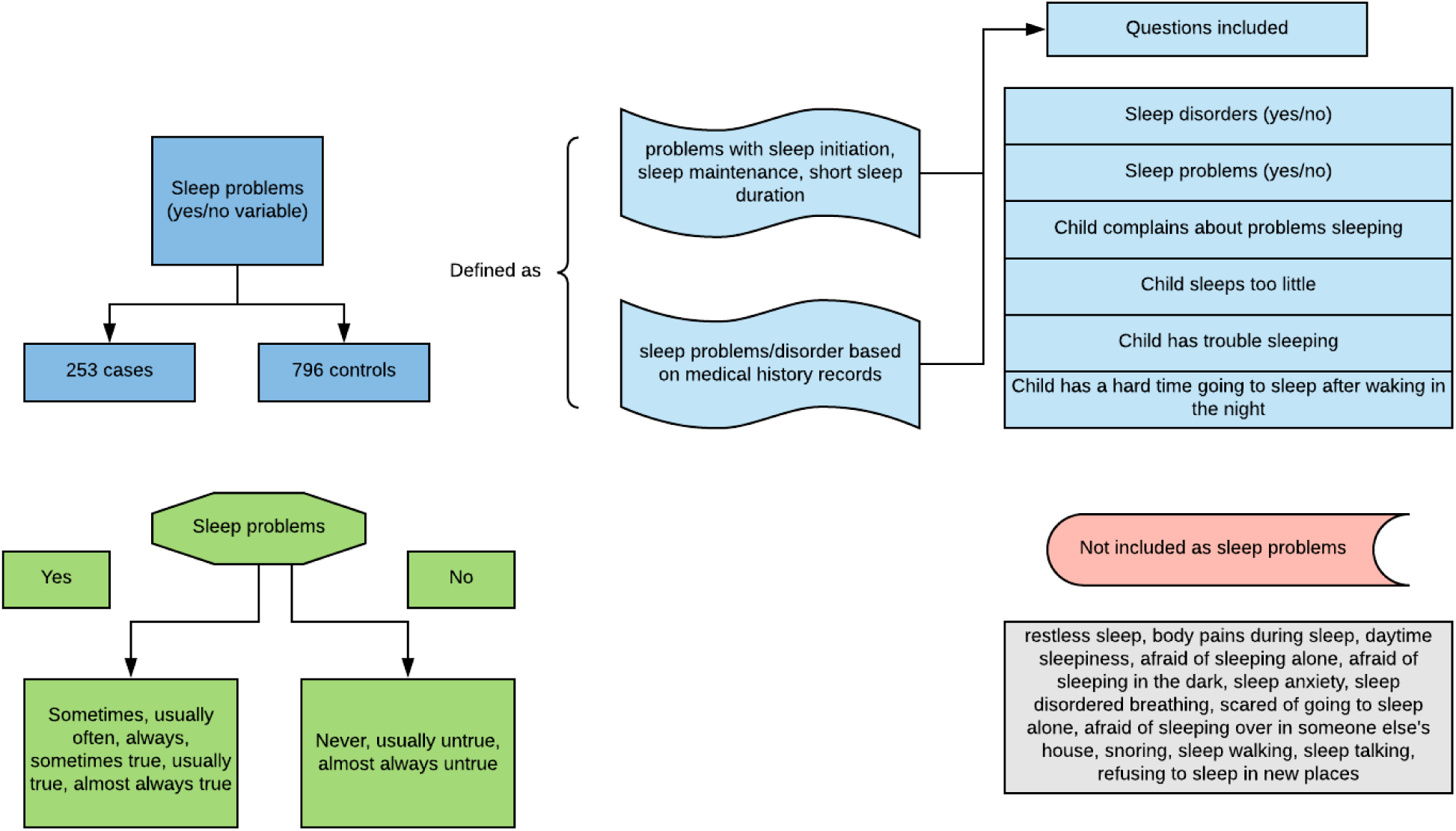
Selection of insomnia cases and controls in the MSSNG dataset

**Figure 3a.**
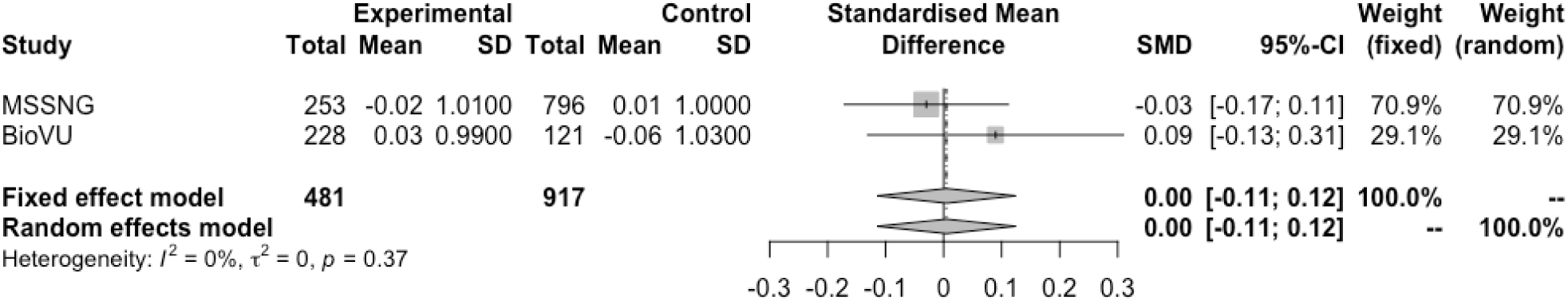
Meta-analyses results of insomnia PRS predicting sleep problems in BioVU and MSSNG

**Figure 3b.**
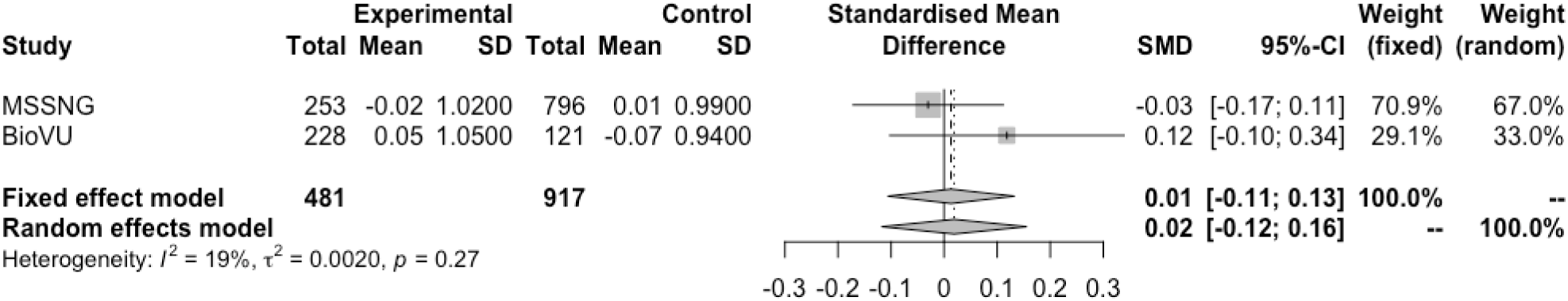
Meta-analyses results of insomnia PRS predicting sleep problems in BioVU and MSSNG

### Statistical Analyses

#### PGS analyses

We constructed PGS of insomnia using the largest GWAS to date^9^, in a total of 349 individuals from the BioVU sample and 1,049 individuals of the MSSNG sample. The PGS per individual was estimated using a continuous shrinkage prior (CS) to SNP effect sizes using the PRS_CS software. We standardized the PGS to have a mean of 0 and a standard deviation of 1. Further, we examined whether the PGS of insomnia was associated with a clinical diagnosis of insomnia within each sample, while also adjusting for sex, age at the time of chart review, and the 10 Principal Components of genetic ancestry. We then meta-analyzed the results between the BioVU sample and the MSSNG sample using the R package ‘meta’^26^. Next, we repeated the aforementioned analyses, but instead of the PGS of insomnia, we constructed PGS trained on the latest GWAS of autism^27^.

#### Sequence Kernel Association Test (SKAT)

We used the Sequence Kernel Association Test (SKAT)^28^ applied to the MSSNG dataset to test whether there is a greater burden of rare variation (Minor Allele Frequency <=0.03) in the melatonin and circadian genes in individuals with autism and insomnia than in individuals with autism and without insomnia. We included age, sex and 10 principal components generated from genetic data as covariates. We also tested whether the combined effects of common and rare variants within the circadian and melatonin genes may be related to insomnia in individuals with autism. Finally, we conducted the same SKAT analyses in the BioVU sample. We then used MetaSKAT^29^ to meta-analyze these results.

#### PrediXcan analyses

We applied PrediXcan^30^ in the BioVU sample, to examine whether the predicted expression of 1) genes related to the circadian rhythms, 2) genes implicated in autism^31^, and 3) genes in the melatonin pathway, are associated with sleep problems. The determination of the tissue that was tested per gene was based on the best prediction performance (r^2^) of the gene on the tissue using the elastic net approach^30^ and the Unified Test for Molecular Signatures (UTMOST) penalized regression model^32^. All gene lists are provided in table 2 and supplementary table 2. We adjusted the tested models for 10 principal components estimated from genetic data.

## Results

### PGS analyses

There were N=253 individuals with insomnia in the MSSNG dataset and N=796 controls while there were N=228 individuals with insomnia in BioVU and N=121 controls. The associations between the PGS of insomnia and sleep problems (i.e., any problem with sleep initiation, sleep maintenance and short sleep duration) in both the BioVU and MSSNG datasets were not significant (b=0.02, standard error (s.e)=0.03, p=0.39 and b=-0.04, s.e.=0.07, p=0.57 respectively). When we meta-analyzed the results from both the BioVU and the MSSNG samples, there was no evidence for heterogeneity between the two studies (p=0.37). There was also no evidence that the insomnia PGS associated with insomnia in the autism sample (random effects model, standard mean difference (SMD)=0.005, p=0.94). Similarly, when we meta-analyzed the results from both the BioVU and the MSSNG samples for the autism PGS, there was no evidence for heterogeneity between the two studies (p=0.27). There was also no evidence of association between the autism PGS and insomnia in the autism sample (random effects model, SMD=0.02, p=0.79)

### SKAT

When we performed the SKAT analyses in the MSSNG dataset, there was nominal evidence of association between rare variants within the Basic Helix-Loop-Helix Family Member E41 (BHLHE41) gene (p=0.03), but it did not survive correction for multiple testing (p<0.01) (Table 1). We also performed the SKAT analyses in BioVU, and again observed weak evidence of association between rare variation in the BHLHE41 gene (p=0.11) and insomnia in the BioVU autism dataset (Table 1). We then used MetaSKAT to meta-analyze these results, and the association with the BHLHE41 gene was no longer significant (p=0.39) (Table 1). We also tested whether the combined effects of common and rare variants within the circadian and melatonin genes may be related to insomnia in individuals with autism, but there was no evidence for associations (Supplementary table 1).

**Table 1.**
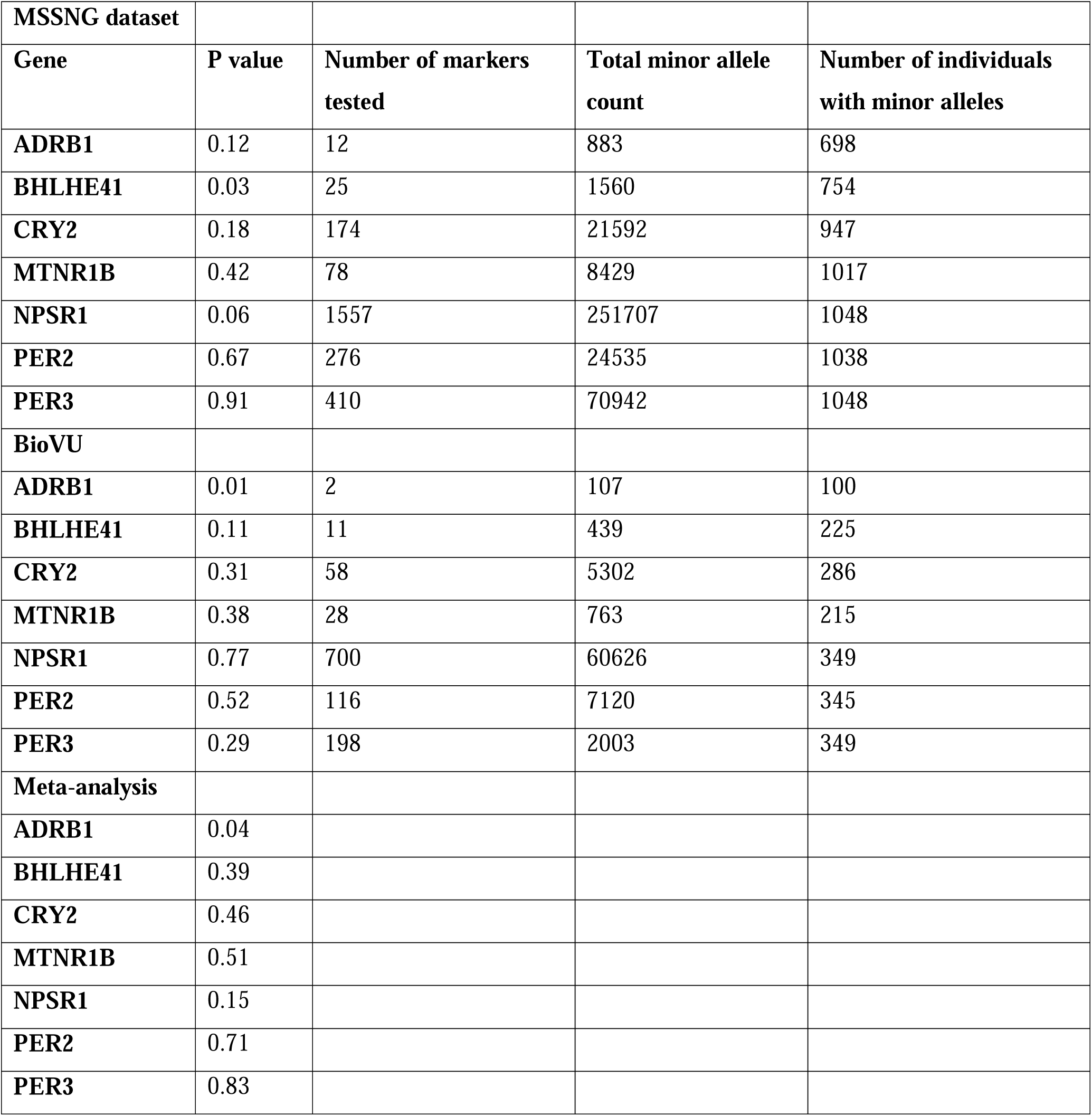
Results from the SKAT analyses

| **MSSNG dataset** |  |  |  |  |
| --- | --- | --- | --- | --- |
| **Gene** | **P value** | **Number of markers tested** | **Total minor allele count** | **Number of individuals with minor alleles** |
| **ADRB1** | 0.12 | 12 | 883 | 698 |
| **BHLHE41** | 0.03 | 25 | 1560 | 754 |
| **CRY2** | 0.18 | 174 | 21592 | 947 |
| **MTNR1B** | 0.42 | 78 | 8429 | 1017 |
| **NPSR1** | 0.06 | 1557 | 251707 | 1048 |
| **PER2** | 0.67 | 276 | 24535 | 1038 |
| **PER3** | 0.91 | 410 | 70942 | 1048 |
| **BioVU** |  |  |  |  |
| **ADRB1** | 0.01 | 2 | 107 | 100 |
| **BHLHE41** | 0.11 | 11 | 439 | 225 |
| **CRY2** | 0.31 | 58 | 5302 | 286 |
| **MTNR1B** | 0.38 | 28 | 763 | 215 |
| **NPSR1** | 0.77 | 700 | 60626 | 349 |
| **PER2** | 0.52 | 116 | 7120 | 345 |
| **PER3** | 0.29 | 198 | 2003 | 349 |
| **Meta-analysis** |  |  |  |  |
| **ADRB1** | 0.04 |  |  |  |
| **BHLHE41** | 0.39 |  |  |  |
| **CRY2** | 0.46 |  |  |  |
| **MTNR1B** | 0.51 |  |  |  |
| **NPSR1** | 0.15 |  |  |  |
| **PER2** | 0.71 |  |  |  |
| **PER3** | 0.83 |  |  |  |

### PrediXcan

We found no evidence that the predicted expression of genes related to circadian rhythms (Table 2), melatonin (Table 2), or autism (Supplementary table 2) are differentially expressed in individuals with autism and insomnia compared to those with autism alone (Bonferroni p-value >0.005). Thanks to our detailed phenotypic efforts in BioVU, we were able to also examine melatonin response in patients with sleep problems and autism who had also been prescribed melatonin supplements. Of those with melatonin supplement use documented in their medical charts, a total of N=46 responded to melatonin treatment and N=53 did not respond to melatonin treatment. There was no evidence that melatonin pathway genes showed differential predicted expression between patients who responded and those who did not respond to melatonin (*MTNR1B*: p=0.55, *AANAT*: p=0.45).

**Table 2.** PrediXcan results in BioVU

| Circadian genes |  |  | Model: sleep problem (yes/no) ~<br>gene +10 PCs |  |
| --- | --- | --- | --- | --- |
|  | p value | OR | lower 95% CI | upper 95% CI |
| **ADRB1** | 0.26 | 1.1 | 0.94 | 1.28 |
| **CRY1** | 0.44 | 1.05 | 0.94 | 1.17 |
| **CRY2** | 0.85 | 1.02 | 0.8 | 1.31 |
| **CSNK1D** | 0.96 | 0.99 | 0.81 | 1.22 |
| **BHLHE41** | 0.93 | 1.01 | 0.87 | 1.16 |
| **NPSR1** | 0.44 | 1.06 | 0.92 | 1.21 |
| **PER2** | 0.63 | 1.12 | 0.71 | 1.77 |
| **PER3** | 0.5 | 0.94 | 0.77 | 1.13 |
| **TIMELESS** | 0.94 | 0.99 | 0.85 | 1.17 |
| Melatonin genes results |  |  | Model: sleep problem (yes/no) ~<br>gene +10 PCs |  |
| **MTNR1B** | 0.56 | 0.94 | 0.78 | 1.15 |
| **AANAT** | 0.2 | 1.52 | 0.93 | 2.49 |
| Melatonin genes results |  |  | Model: melatonin<br>response(yes/no)~ gene +10 PCs |  |
| **MTNR1B** | 0.55 | 0.84 | 0.48 | 1.47 |
| **AANAT** | 0.45 | 1.87 | 0.38 | 9.29 |

## Discussion

This is the first study to test whether polygenic scores of insomnia or autism were related to risk of insomnia in people with autism, and whether the circadian genes were associated with insomnia in people with autism. To answer these questions, we employed the phenotypically and genotypically unique MSSNG dataset, that is enriched for individuals with autism. We also incorporated data from the Vanderbilt University Biobank (BioVU) including validated autism diagnoses as well as carefully curated phenotyping on insomnia in autism patients. Overall, we did not find evidence for strong effects of genetic scores influencing sleep in people with autism, however, we cannot rule out the possibility that genetic factors may play a smaller role in sleep problems, which we were underpowered to detect.

Specifically, we constructed PGS of insomnia and autism in both samples and tested their associations with insomnia in individuals with autism. In our meta-analyzed sample, no associations reached statistical significance. To take advantage of the rare variant data available in MSSNG, we also tested whether there is a greater burden of rare and disruptive variation in the melatonin and circadian genes in individuals with autism and insomnia than in individuals with autism and without insomnia. There was weak evidence that the Basic Helix-loophelix family member e41 (*BHLHE41*) gene was associated with insomnia in people with autism from the MSSNG sample, although this association did not survive correction for multiple testing.

This gene has been previously associated with short sleep duration^33^ and appears to regulate sleep length in mammals^34^. There was no evidence for any other associations. Moreover, there was no evidence to suggest that the predicted expression of genes related to melatonin, circadian rhythms and autism were related to insomnia.

Although we used the largest sample sizes available in combination with advanced genotyping and phenotyping methods, the lack of associations between the PGS of insomnia and insomnia in the individuals with autism is likely due to limited power. It is also possible that the genetics of insomnia in the general population are different than the genetics of insomnia in individuals with autism, however, the polygenicity of insomnia and autism and the variants of small effect sizes associated with both traits points to the need for a larger collection of data on sleep problems and sleep quality among people with autism. Similarly, the lack of associations with predicted expression, as well as lack of evidence for a greater burden of rare variation in the melatonin and circadian genes, could also be due to a Type 2 error.

Finally, the small effect sizes of the genetic variants associated with autism and insomnia, may also indicate that the sleep problems experienced by people with autism may be more related to a variety of causes, including higher level system biology. For instance, they may be a function of sensitivity to the environment (e.g., noise, or thermal comfort)^35^ or other environmental triggers. They may also be related to medical causes, such as gastrointestinal disease, medications (e.g., stimulants for attention deficit-hyperactivity disorder or antidepressants for anxiety/depression), or behavioral factors such as excessive screen time or limited exercise^1^. These factors may be more readily modifiable and future research is necessary to establish their role in the sleep problems of people with autism.

## Data Availability

Regarding the BioVU sample, due to data sharing restrictions related to privacy concerns in the EHR, the datasets generated from our hospital population will not be publicly available. Regarding the MSSNG sample, data are available in the MSSNG repository (http://research.mss.ng)

## Acknowledgements

We would like to thank the participating families and the resources of MSSNG and Autism Genetic Resource Exchange (AGRE) as well as Autism Speaks. We would also like to thank the Synthetic Derivative: The project described was supported by the National Center for Research Resources, Grant UL1 RR024975-01, and is now at the National Center for Advancing Translational Sciences, Grant 2 UL1 TR000445-06. The content is solely the responsibility of the authors and does not necessarily represent the official views of the NIH.

## Notes

### Competing Interest Statement

The authors have declared no competing interest.

### Funding Statement

The study was funded by Autism Speaks (#11680)

